# Clinical and economic benefits of lenzilumab plus standard of care compared with standard of care alone for the treatment of hospitalized patients with Coronavirus Disease 19 (COVID-19) from the perspective of National Health Service England

**DOI:** 10.1101/2022.02.11.22270859

**Authors:** Adrian Kilcoyne, Edward Jordan, Kimberly Thomas, Alicia N. Pepper, Allen Zhou, Dale Chappell, Miyuru Amarapala, Rachel-Karson Thériault, Melissa Thompson

**Affiliations:** HEOR, Humanigen Inc, Burlingame, California, United States; Value & Evidence Services, EVERSANA, Burlington, Ontario, Canada

**Keywords:** economic analysis, CRP, GM-CSF, ventilator-free survival, survival without ventilation, invasive mechanical ventilation

## Abstract

**Purpose:** Estimate the clinical and economic benefits of lenzilumab plus standard of care (SOC) compared with SOC alone in the treatment of hospitalized COVID-19 patients from the National Health Service (NHS) England perspective.

**Methods:** A cost calculator was developed to estimate the clinical benefits and costs of adding lenzilumab to SOC in newly hospitalized COVID-19 patients over 28 days. The LIVE-AIR trial results informed the clinical inputs: failure to achieve survival without ventilation (SWOV), mortality, time to recovery, intensive care unit (ICU) admission, and invasive mechanical ventilation (IMV) use. Base case costs included drug acquisition and administration for lenzilumab and remdesivir and hospital resource costs based on level of care required. Clinical and economic benefits per weekly cohort of newly hospitalized patients were also estimated.

**Results:** In all populations examined, specified clinical outcomes were improved with lenzilumab plus SOC over SOC treatment alone. In a base case population aged <85 years with C-reactive protein (CRP) <150 mg/L, with or without remdesivir, adding lenzilumab to SOC was estimated to result in per-patient cost savings of £1,162. In a weekly cohort of 4,754 newly hospitalized patients, addition of lenzilumab to SOC could result in 599 IMV uses avoided, 352 additional lives saved, and over £5.5 million in cost savings. Scenario results for per-patient cost savings included: 1) aged <85 years, CRP <150 mg/L, and receiving remdesivir (£3,127); 2) Black patients with CRP <150 mg/L (£9,977); and 3) Black patients from the full population (£2,369). Conversely, in the full mITT population, results estimated additional cost of £4,005 per patient.

**Conclusion:** Findings support clinical benefits for SWOV, mortality, time to recovery, time in ICU, time on IMV, and ventilator use, and an economic benefit from the NHS England perspective when adding lenzilumab to SOC for hospitalized COVID-19 patients.

## Introduction

The societal and economic impacts of the Coronavirus Disease 2019 (COVID-19) pandemic have been substantial in the United Kingdom (UK) and worldwide.^1–3^ As of January 25, 2022, the World Health Organization has reported over 352 million COVID-19 cases and over 5.6 million deaths globally.^4^ As of January 25, 2022, over 16 million people in the UK have tested positive for COVID-19, with 685,624 patients being admitted to hospitals and 176,813 deaths with COVID-19 on the death certificate.^5–7^ A study of 20,133 patients hospitalized with COVID-19 in the UK found that 17% of hospitalized patients required admission to the intensive care unit (ICU) and of these patients, 32% died.^8^ Moreover, 10% of patients hospitalized for COVID-19 required invasive mechanical ventilation (IMV), with 37% of these patients dying.^8^

Despite approximately 83.9% and 64.3% of the UK population aged ≥12 years having received two and three vaccination doses, respectively, as of January 25, 2022,^9^ the need for effective treatments for hospitalized COVID-19 patients remains, particularly with the continued emergence of new COVID-19 variants, including the Omicron variant in November 2021.^10^ Whilst there is evidence of vaccine effectiveness against symptomatic infection and hospitalization, a highly transmissible variant such as Omicron can lead to an increase in hospital admissions.^11,12^ As of January 23, 2022, the number of COVID-19 patients to admitted UK hospitals in the past 30 days more than doubled, relative to the previous 30-day period.^13^ As new variants continue to emerge, there remains an urgent unmet need for effective therapies for patients who are hospitalized with COVID-19 to prevent IMV and/or death.

There is a substantial economic burden associated with the treatment of COVID-19 in terms of overall healthcare costs.^14,15^ From the hospital perspective, providing care to patients with COVID-19 in the UK is associated with average per-patient daily costs ranging from a low of 883 euros (EUR) for patients who do not require ICU admission or IMV to a high of 3,183 EUR for patients who require both ICU admission and IMV (as reported in 2020).^16^ Given that hospitalized patients with COVID-19, especially those requiring IMV, have more detrimental health outcomes and a higher economic burden, safe and effective treatment options that provide good value for money represent a significant unmet need.^17–19^

An increased risk of IMV and death in COVID-19 patients has been linked to a process known as immune-mediated hyperinflammation.^20^ This process is largely mediated by granulocyte-macrophage colony-stimulating factor (GM-CSF), a protein that is produced locally in inflamed tissue in response to COVID-19 infection.^21,22^ Activated GM-CSF functions to activate and mobilize myeloid cells, resulting in the excessive production of downstream pro-inflammatory cytokines, including interleukin-6 (IL-6).^21,23,24^ C-reactive protein (CRP) is an acute phase reactant produced by the liver largely in response to IL-6, and CRP levels serve as a reliable surrogate for IL-6 bioactivity.^25,26^ Lung tissue injury, disease severity, and ICU admission of patients with COVID-19 have been directly correlated with both GM-CSF levels^27–30^ and CRP levels at the time of hospital admission.^31–37^ Further, neutralization of GM-CSF is expected to prevent or reduce the severity and sequelae of immune-mediated hyperinflammation.^24^ As such, GM-CSF has been identified as an important therapeutic target of COVID-19.^29^

Lenzilumab is a novel Humaneered^®^ anti-human GM-CSF monoclonal antibody that directly binds GM-CSF, thereby preventing its downstream signaling.^23^ This mechanism of action of lenzilumab renders it a promising new treatment option as its efficacy is likely unaffected by new COVID-19 variants.^38^ LIVE-AIR (NCT04351152) was a Phase 3, prospective, randomized, multicentered, double-blind, placebo-controlled clinical trial that evaluated the efficacy and safety of lenzilumab or placebo (in addition to institutional standard of care [SOC] in both treatment groups) in hospitalized participants with COVID-19 pneumonia.^23^ Patients must have been hospitalized with an oxygen saturation (SpO_2_) ≤94% on room air or required supplemental oxygen but had not progressed to IMV.^23^ Results from the LIVE-AIR trial showed that lenzilumab plus SOC significantly improved the likelihood of achieving survival without ventilation (SWOV)^a^ (sometimes referred to as ventilator-free survival) by day 28 compared with placebo plus SOC.^23^ Additional analyses revealed that lenzilumab plus SOC had greater efficacy in patients aged <85 years with CRP levels <150 mg/L, overall and exclusively in patients receiving remdesivir, compared with placebo plus SOC, as measured by SWOV.^39,40^ As previously mentioned, CRP levels are elevated in the process of immune-mediated hyperinflammation and have been linked to poor clinical outcomes among patients with severe COVID-19.^23,34,35,40^ Therefore, evidence from the LIVE-AIR trial suggest that lenzilumab may have greater efficacy when administered as an early intervention in hospitalized patients with COVID-19.

Results from an exploratory analysis of the LIVE-AIR trial also suggested that Black and African American patients with baseline level CRP <150 mg/L may exhibit the greatest response to lenzilumab, with a nearly 9-fold increase in the likelihood of SWOV.^41^ This is notable as the Office of National Statistics (ONS) reported that the COVID-19 death rate was higher for Black African (2.16 greater for males and 1.62 greater for females) and Black Caribbean (1.69 greater for males and 1.35 greater for females) populations in the UK compared to their White British counterparts, during the second wave of the COVID-19 pandemic (September 12, 2020 to March 31, 2021).^42^ The hyper-vulnerability in this population can be largely attributed to low vaccination rates, as a recent UK-wide survey identified Black or Black British respondents as having the highest rate of vaccine hesitancy (71.8%).^43^ Further, the Black population, on average, has a higher incidence and prevalence of chronic illness (eg, diabetes, hypertension, obesity) and lower socioeconomic status, both of which result in higher risk of infection, hospitalization, and death.^43,44^

Although clinical evidence supports that lenzilumab effectively improves the likelihood of SWOV in hospitalized patients with COVID-19,^23,40^ its overall clinical and economic value from the National Health Service (NHS) England perspective has not been previously characterized. The objective of this study was to report the clinical benefits of the LIVE-AIR trial in a meaningful way to the NHS England perspective, and to estimate the per-patient and population-level costs using the clinical trial data to demonstrate the economic benefit of lenzilumab plus SOC compared with SOC alone in the treatment of hospitalized COVID-19 patients. This analysis was conducted *ex ante* (ie, before regulatory approval of lenzilumab).

## Methods

### Calculator Structure

We previously developed a Microsoft^®^ Excel-based cost calculator to estimate the clinical benefits and per-patient costs of lenzilumab plus SOC versus SOC alone for newly hospitalized patients with COVID-19 pneumonia from a United States (US) hospital perspective.^45^ This calculator was adapted to represent a UK healthcare payer perspective, specifically that of NHS England. A 28-day model time horizon for the index COVID-19 hospitalization was selected to align with data censoring for the LIVE-AIR trial (28 days following enrollment).^23^ A patient-level analysis was conducted to estimate the clinical benefits and per-patient costs for an average of newly hospitalized patients with COVID-19 pneumonia in England. A cohort-level analysis was also conducted considering all newly hospitalized patients in a 1-week period. The calculator structure is presented in **Figure 1**.

**Figure 1.**
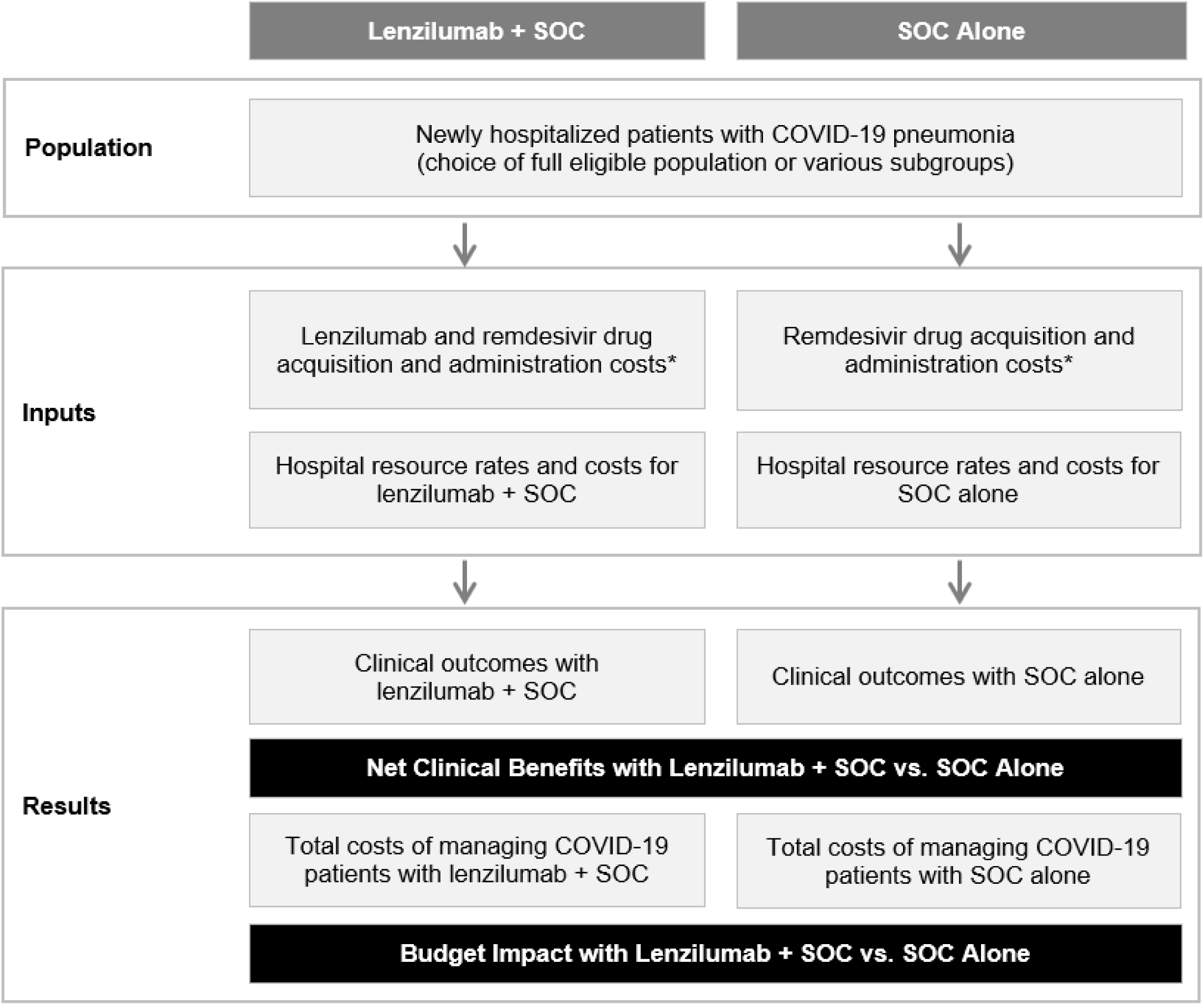
Cost calculator structure^a^ **Note** ^a^ It was assumed that use of SOC drugs, other than remdesivir, would not be affected by the concomitant use of lenzilumab, resulting in no cost differences for SOC drugs between both groups. This assumption was supported by the balanced use of corticosteroids in both treatment arms of the LIVE-AIR trial.^23^ **Abbreviations:** COVID-19, Coronavirus Disease 19; SOC, standard of care.

### Target Population

Patient eligibility for the calculator was based on inclusion criteria for the LIVE-AIR trial. In brief, eligible patients were newly hospitalized with COVID-19 pneumonia, with SpO^2^ ≤94% on room air and/or requiring supplemental oxygen, but not on IMV.^23^ Further analyses of LIVE-AIR trial data suggested that patients aged <85 years with CRP <150 mg/L particularly benefited from lenzilumab, and this population was selected for the base case.^39,40^ Since the ongoing and fully enrolled ACTIV-5/BET-B trial, which aims to further elucidate the efficacy of lenzilumab in COVID-19 in patients aged <85 years with CRP <150 mg/L and receiving remdesivir, this population was explored in a scenario analysis (scenario #1).^46,47^ This population receiving remdesivir was not selected for the base case given the lower use of remdesivir in the UK (16.7%^48^ to 48.4%^49^) compared with that observed in the LIVE-AIR trial (72.4%) conducted primarily in the US.^23^ Scenario analyses were also conducted in the full LIVE-AIR modified intent-to-treat (mITT) population^b^ (scenario #2), in Black patients with CRP <150 mg/L (with or without remdesivir) (scenario #3), and in Black patients from the full mITT population (scenario #4). Data for the mITT population of the LIVE-AIR trial, the pre-specified primary analysis population, were used for all analyses.^50^

### Cohort Size Estimates

Weekly cohort sizes for the base case and scenario analyses were calculated using recent case count reports from UK government sources, the LIVE-AIR trial, and a physician survey. The physician survey was conducted between August 13 to 25, 2021 with 82 medically qualified and practicing pulmonary medicine physicians, infectious disease specialists, and intensive care specialists in the UK.^49^ All physicians were consultants and senior non-consultant hospital doctors and all had treated patients with COVID-19.^49^ In a 1-week period from January 17 to January 23, 2022, there were an estimated 11,343 newly hospitalized COVID-19 patients in England.^13^ Based on the physician survey, 60.1% of hospitalized patients have SpO_2_ less than 94%,^49^ and would be eligible for treatment in the base case and scenario analyses. To calculate all subgroups with CRP <150 mg/L (base case, scenario #1 and #3), it was assumed that 77.9% of the population with SpO_2_ <94% had CRP <150 mg/L, as observed in the LIVE-AIR trial population.^40^ For subgroups restricted to age <85 years (base case, scenario #1), it was assumed that 89.5% of the hospitalized population was <85 years according to NHS data for hospital admissions in 2020-2021.^51^ The subgroup receiving remdesivir (scenario #1) was calculated using data from the physician survey for the percentage of patients with CRP <150 mg/L who are treated with remdesivir, which was estimated at 37.7%.^49^ Finally, for the scenarios in the Black population (scenario #3 and #4) it was assumed that 3.8% of patients were Black according to NHS hospital admissions in 2020-2021.^51^ This resulted in a weekly cohort size of eligible patients of 4,754 for the base case and cohort sizes of 1,792, 6,817, 200, and 256 patients for scenarios #1 through 4, respectively. The calculations for the cohort size estimates are illustrate in **Figure 2**.

**Figure 2.**
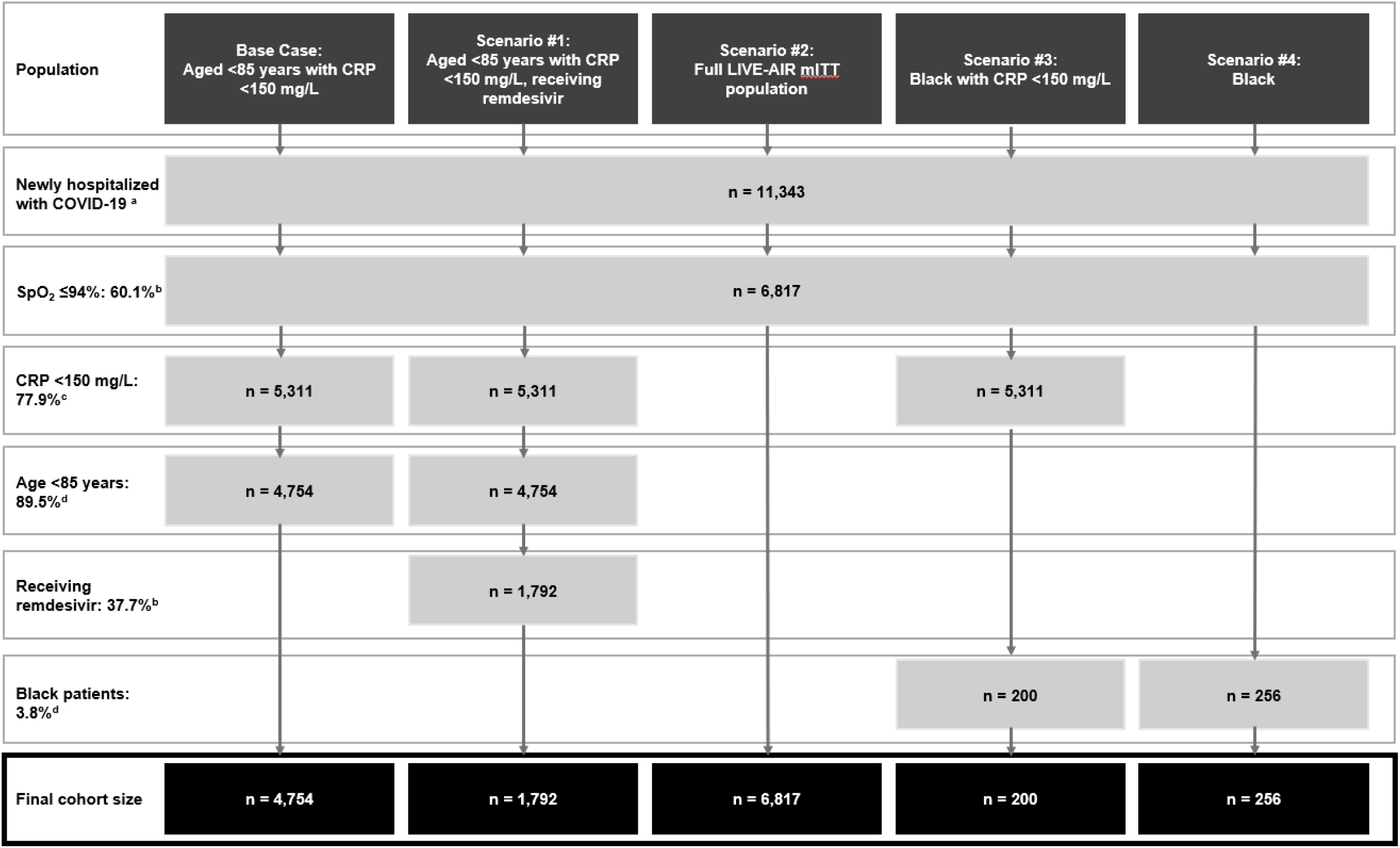
Calculations of the weekly cohort size estimates^a^ **Notes** ^a^ Estimated cohort sizes were calculated using the total number of new COVID-19 hospital admissions in England over a one-week period (11,343; from January 17 to January 23, 2022).^13^ Based on the physician survey, 60.1% of hospitalized patients have SpO_2_ ≤ 94%^49^ and it was assumed that 77.9% of this population has CRP <150 mg/L, as observed in the LIVE-AIR trial.^23^ For base case and scenario #1, it was assumed that 89.5% of the hospitalized population was <85 years^51^ and the subgroup receiving remdesivir was calculated using data from the physician survey which estimated 37.7%.^49^ Lastly, for scenarios #3 and #4, it was assumed that 3.8% of the patient were Black.^51^ ^b^ Source: Government of the United Kingdom.^13 c^ Source: Humanigen. Data on file.^49^ ^d^ Source: Temesgen et al. (2021).^23^ ^e^ Source: National Health Service Digital.^51^ **Abbreviations:** COVID-19, Coronavirus Disease 19; CRP, C-reactive protein; mITT, modified intent-to-treat; SpO_2_, oxygen saturation.

### Treatment Efficacy

Calculator inputs for treatment efficacy of lenzilumab plus SOC versus SOC alone were obtained from the LIVE-AIR trial.^50^ Treatment efficacy inputs for the base case and scenario analyses included failure to achieve SWOV, mortality, time to recovery, time in ICU, and time on IMV, as previously reported in the US hospital perspective analysis by Kilcoyne and colleagues (2022).^45^ Mean time to recovery^c^ was used as a proxy for a patient’s length of stay. The values used for the treatment efficacy inputs are provided in the **Supplementary Appendix**.

### Clinical Outcomes

Using the treatment efficacy data from the LIVE-AIR trial, clinical outcomes were then reported in a manner relevant to healthcare payers. This included number needed to treat (NNT) for one patient to achieve SWOV, reduction in IMV use, NNT for one life saved, and the number of bed days, ICU days, and IMV days saved.

#### Cost Calculator

Base case costs included drug acquisition and administration for lenzilumab and remdesivir and hospital resource costs based on the level of care required. It was important to consider the proportion of patients in each treatment arm expected to receive remdesivir given the difference in the use in UK clinical practice compared with that observed in the LIVE-AIR trial. Additional details on the assumptions and calculations for remdesivir use can be found in the **Supplementary Appendix**.

Adverse event (AE) management costs were not considered because minimal differences in the incidence of grade ≥3 AEs were observed in the LIVE-AIR trial (under 3% difference for all individual AEs) and the overall incidence of grade ≥3 AEs was lower in the lenzilumab plus SOC arm compared to the placebo plus SOC treatment arm (26.7% versus 32.7% for the safety populations, respectively).^23^

### Drug Acquisition Costs

A targeted literature review was conducted to support costing parameterization for the calculator. All costs were reported in 2021 pound sterling (GBP), with adjustments for inflation made as needed based on the consumer price inflation (CPI) time series (CPI INDEX 06: HEALTH) from the ONS.^52^ It was assumed that 72.1% of lenzilumab plus SOC patients received concomitant remdesivir in the base case and all scenario analyses (excluding scenario #1), in alignment with the actual use in the LIVE-AIR trial.^23^ In the base case analysis and Scenario #3 (Black with CRP <150 mg/L), remdesivir use was estimated at 37.7% for patients who received SOC alone, based on the physician survey for treatment of patients with CRP <150 mg/L.^49^ As scenario #1 considered patients aged <85 years with CRP <150 mg/L and receiving remdesivir, remdesivir use was set at 100% for both the lenzilumab plus SOC and SOC alone arms. For scenarios #2 (full mITT population) and #4 (Black patients from the full mITT population), 48.4% of patients in the SOC alone arm were assumed to receive remdesivir, based on the physician survey for treatment of patients with SpO_2_ ≤94%, regardless of CRP level.^49^ Additional details on the assumptions and calculations for remdesivir use can be found in the **Supplementary Appendix**.

The drug acquisition cost for lenzilumab was £7,300 per patient for the entire treatment course, based on the official UK list price;^53^ this was calculated as a cost of £406 per 100 mg vial and a dosing regimen of three 600 mg doses, each administered by a 1-hour intravenous infusion 8 hours apart, for a total of 1800 mg per patient.^23^ The total drug acquisition cost for remdesivir was £2,040; this was calculated using a cost of £340 per 100 mg vial^54^ and assuming a course of 6 vials based on the recommended 200 mg loading dose on day 1 and 100 mg maintenance doses on days 2-5.^55^

### Drug Administration Costs

Lenzilumab administration costs were assumed to be £33.92 per intravenous infusion, for a total administration cost of £101.76. This cost was calculated based on the pharmacy labor cost associated with the aseptic preparation and administration of a monoclonal antibody.^56,57^ Aseptic technique was included for lenzilumab as its preparation requires the compounding of six 100 mg vials and is therefore considered a medium-risk for contamination.^58^ It was assumed that remdesivir administration costs were £14.75 per infusion, for a total administration cost of £73.75. This cost was calculated based on the pharmacy labor cost associated with administration of a monoclonal antibody, with no aseptic preparation.^57^ It was assumed that additional time for aseptic compounding was not needed for remdesivir as its preparation includes combination of no more than two drug vials and thus is considered a low-risk for contamination.^58^

Nursing administration costs for intravenous infusions, as well as drug and administration costs for other SOC drugs, including corticosteroids, were assumed to be captured in the hospital resource costs. Costs associated with other SOC drugs were not included as separate inputs in the calculator because it was assumed that lenzilumab would not impact the utilization or cost of background therapies other than remdesivir; this assumption was supported by balanced corticosteroid use in both arms of the LIVE-AIR trial.^23^ Additional details on calculations for administration costs can be found in the **Supplementary Appendix**.

### Hospital Resource Use and Costs

In alignment with the US hospital perspective model,^45^ patients were divided into four levels of care required during their hospital stay: no ICU and no IMV, ICU but no IMV, IMV but no ICU, and both ICU and IMV. Daily hospital resource cost inputs were obtained from a study by Czernichow and colleagues (2020), which reported mean cost per bed type per day in the UK.^16^ A cost corresponding to the “IMV but no ICU” level of care was not provided in the study, so it was assumed to be equivalent to the difference between the “both ICU and IMV” and “ICU but no IMV” groups, added to the base hospitalization cost (ie, “no ICU or IMV”). Czernichow and colleagues reported all costs in their study in EUR, using an exchange rate of 1.11 EUR to 1 GBP;^16^ these costs were reconverted using that exchange rate and then inflated from 2020 to 2021 values to provide the following hospital costs per day: £876 (no ICU, no IMV), £1,978 (ICU, but no IMV), £2,043 (IMV, but no ICU), and £3,145 (both ICU and IMV).^52^ It was assumed that these daily costs incorporated all costs associated with SOC in the UK, including any corticosteroid use. Inputs for hospital resource use were informed by data from the LIVE-AIR trial^50^ and included patient distributions and time to recovery for each level of care, as previously reported by Kilcoyne and colleagues (2022).^45^ Costs and resource use for each level of care are provided in the **Supplementary Appendix**.

Hospital resource costs per patient were calculated as a weighted average based on the four levels of care required. First, the average time to recovery for each of the four levels of care was multiplied by the corresponding daily hospital resource cost to obtain the average total hospital resource cost for each level of care. Next, the weighted average cost per patient was calculated as the sum product of average total hospital resource costs for each level of care and the corresponding proportion of patients requiring each level of care. These calculations were performed independently for the lenzilumab plus SOC and SOC alone arms to obtain separate weighted average hospital resource costs for the two treatment options. Additional details on hospital resource cost calculations are provided in the **Supplementary Appendix**.

### Total Costs and Economic Impact

Total costs were calculated for each treatment arm; this included drug costs, administration costs, and hospital resource costs. The difference in total costs between the two arms was then calculated to estimate the average cost per patient treated with lenzilumab plus SOC instead of SOC alone. To obtain results for the weekly cohort, the average cost per patient was multiplied by the number of newly hospitalized patients eligible for treatment in a 1-week period.

#### Scenario Analyses

The base case analysis included patients aged <85 years with CRP <150 mg/L. Four scenario analyses were conducted to examine the per-patient cost of adding lenzilumab to SOC in other patient populations. Scenario analysis #1 included a narrower population by only including base case patients who received concomitant remdesivir. Scenario analysis #2 examined a broader population and included the full LIVE-AIR mITT population. The other two scenario analyses focused on Black patients, a population that is disproportionally affected by COVID-19.^42–44^ Scenario analysis #3 included Black patients with CRP <150 mg/L, with or without remdesivir, whereas scenario analysis #4 included Black patients from the full mITT population.

#### Sensitivity Analyses

Sensitivity analyses were conducted to determine the key drivers of the cost calculator. The following calculator inputs were included for sensitivity analyses: daily hospital resource costs, patient distributions in each level of care (lenzilumab plus SOC arm only), lenzilumab drug costs, and remdesivir use (SOC alone arm only). Daily hospital resource cost inputs were adjusted by ±25% and each level of care was adjusted simultaneously. Given that in the cost calculator, the total hospital resource cost at each level of care is the product of the respective daily hospital resource cost and the time to recovery, the adjustment of daily hospital resource costs by ±25% has the same impact as adjusting time to recovery by ±25%; therefore, additional sensitivity analyses for time to recovery were not conducted. For patient distributions, the proportion of patients requiring both ICU and IMV was adjusted ±25% first and then the three other levels of care were reweighted, maintaining the proportionality between the levels of care prior to adjustment. The decision to use the ICU and IMV group for adjustment in the sensitivity analysis was based on the assumption that this group was the largest driver of costs in the model, as a result of the increased cost of care per day and the longer length of stay. The input for lenzilumab drug cost was adjusted by +10% and –25%. The variability in the upper estimate (+10% price increase) was selected to be smaller than the lower estimate (−25% price reduction) as the drug cost used in the base case analysis of the cost calculator represents the official UK list price of lenzilumab and, therefore, an increase of more than 10% of the list price in the UK is not anticipated. Lastly, for remdesivir use in the SOC arm, 16.7% was used as the low estimate based on a study by Arch and colleagues^48^ and 72.8% was used as the high estimate based on the placebo plus SOC arm in the LIVE-AIR trial.^23^

## Results

### Base Case and Scenario Analyses

In the base case and all scenario analyses, all specified clinical outcomes were improved with lenzilumab plus SOC over SOC alone (**Table 1**). In the base case, the NNT findings showed that every eighth patient treated with lenzilumab plus SOC rather than SOC alone avoided IMV or death. Across the scenario analyses this NNT ranged from a low of 4 in the Black with CRP <150 mg/L subgroup up to 15 in the full LIVE-AIR mITT population. Lenzilumab also provided a benefit in terms of mortality, with a base case NNT of 14 to save one life and this ranged from 8 to 23 across the scenario analyses. The addition of lenzilumab to SOC compared with SOC alone also reduced ventilator use by 12.6% (ranging from 6.5% to 26.8%) and 3.33 IMV days were saved in the base case analysis (ranging from 1.85 up to 5.50 days). Finally, the time to recovery was improved with lenzilumab plus SOC, saving 2.40 bed days (ranging from 0.99 up to 4.92 days) as well as reducing time in ICU by 2.73 ICU days (ranging from 1.21 up to 5.06 days).

**Table 1.** Estimated clinical benefits of lenzilumab plus SOC over SOC alone per treated patient.

|  | **Base case: aged**<br>**<85 years with**<br>**CRP <150 mg/L** | **Scenario #1:**<br>**aged <85 years**<br>**with CRP <150**<br>**mg/L, receiving**<br>**remdesivir** | **Scenario #2: full**<br>**LIVE-AIR mITT**<br>**population** | **Scenario #3:**<br>**Black with CRP**<br>**<150 mg/L** | **Scenario #4:**<br>**Black** |
| --- | --- | --- | --- | --- | --- |
| **Failure to achieve SWOV<sup>a</sup>** |  |  |  |  |  |
| Lenzilumab plus SOC | 8.4% | 9.2% | 15.6% | 4.0% | 13.2% |
| SOC alone | 21.2% | 24.8% | 22.1% | 29.3% | 29.1% |
| NNT for one patient to achieve SWOV | **8** | **6** | **15** | **4** | **6** |
| **Ventilator use** |  |  |  |  |  |
| Lenzilumab plus SOC | 8.2% | 8.9% | 15.3% | 4.0% | 13.2% |
| SOC alone | 20.8% | 24.4% | 21.8% | 30.8% | 30.3% |
| Reduced IMV use | **12.6%** | **15.5%** | **6.5%** | **26.8%** | **17.1%** |
| **Mortality<sup>a</sup>** |  |  |  |  |  |
| Lenzilumab plus SOC | 6.5% | 7.5% | 9.6% | 4.0% | 7.9% |
| SOC alone | 13.9% | 17.3% | 13.9% | 17.1% | 16.5% |
| NNT for one life saved | **14** | **10** | **23** | **8** | **12** |
| **Time to recovery (days)** |  |  |  |  |  |
| Lenzilumab plus SOC | 9.21 | 9.93 | 11.09 | 8.32 | 10.87 |
| SOC alone | 11.61 | 12.33 | 12.08 | 13.24 | 13.45 |
| Bed days saved | **2.40** | **2.40** | **0.99** | **4.92** | **2.58** |
| **Time in ICU (days)** |  |  |  |  |  |
| Lenzilumab plus SOC | 3.48 | 3.79 | 5.40 | 1.48 | 4.68 |
| SOC alone | 6.21 | 6.76 | 6.61 | 6.54 | 7.06 |
| ICU days saved | **2.73** | **2.97** | **1.21** | **5.06** | **2.38** |
| **Time on IMV (days)** |  |  |  |  |  |
| Lenzilumab plus SOC | 1.92 | 2.12 | 3.53 | 1.12 | 3.11 |
| SOC alone | 5.25 | 6.02 | 5.38 | 6.62 | 6.48 |
| IMV days saved | **3.33** | **3.90** | **1.85** | **5.50** | **3.37** |

**Note:**
<sup>a</sup> These values were derived from Kaplan–Meier analysis.
**Abbreviations:** CRP, C-reactive protein; ICU, intensive care unit; IMV, invasive mechanical ventilation; mITT, modified intent-to-treat; NNT, number needed to treat; SOC, standard of care; SWOV, survival without ventilation.

In addition to improved clinical outcomes, an estimated cost savings of £1,162 per patient resulted from adding lenzilumab to SOC in the base case analysis (**Table 2**). Scenario analysis #1 (aged <85 years with CRP <150 mg/L, with remdesivir) also produced an estimated cost savings of £3,127 per patient. In scenario analysis #2 (full LIVE-AIR mITT population), addition of lenzilumab to SOC was estimated to result in an additional cost of £4,005 per patient over SOC alone. Scenario analysis #3 (Black patients with CRP <150 mg/L, with or without remdesivir) and scenario analysis #4 (Black patients from the full mITT population) estimated cost savings of £9,977 and £2,369 per patient, respectively.

**Table 2.** Estimated economic impact of lenzilumab plus SOC versus SOC alone per treated patient.

|  | Lenzilumab<br>Acquisition<br>Costs | Remdesivir<br>Acquisition<br>Costs<sup>a</sup> | Drug<br>Administration<br>Costs | Hospital<br>Resource Use<br>Costs | Total<br>Treatment<br>Costs | Cost<br>Difference<sup>b</sup> |
| --- | --- | --- | --- | --- | --- | --- |
| Base case: aged <85 years with CRP <150 mg/L |  |  |  |  |  |  |
| Lenzilumab plus SOC | £7,300 | £1,471 | £155 | £15,493 | £24,419 | -£1,162 |
| SOC alone | £0 | £769 | £28 | £24,784 | £25,581 |  |
| Scenario #1: aged <85 years with CRP <150 mg/L, receiving remdesivir |  |  |  |  |  |  |
| Lenzilumab plus SOC | £7,300 | £2,040 | £176 | £16,611 | £26,126 | -£3,127 |
| SOC alone | £0 | £2,040 | £74 | £27,140 | £29,254 |  |
| Scenario #2: full LIVE-AIR mITT population |  |  |  |  |  |  |
| Lenzilumab plus SOC | £7,300 | £1,471 | £155 | £21,824 | £30,750 | £4,005 |
| SOC alone | £0 | £987 | £36 | £25,721 | £26,744 |  |
| Scenario #3: Black with CRP <150 mg/L |  |  |  |  |  |  |
| Lenzilumab plus SOC | £7,300 | £1,471 | £155 | £11,682 | £20,608 | -£9,977 |
| SOC alone | £0 | £769 | £28 | £29,788 | £30,585 |  |
| Scenario #4: Black |  |  |  |  |  |  |
| Lenzilumab plus SOC | £7,300 | £1,471 | £155 | £20,436 | £29,362 | -£2,369 |
| SOC alone | £0 | £987 | £36 | £30,708 | £31,731 |  |

**Note:**
<sup>a</sup> Additional details regarding the assumptions and calculations for remdesivir use can be found in the **Drug Acquisitions Costs** section and in the associated **Supplementary Appendix**.
<sup>b</sup> Negative numbers indicate a net cost savings with lenzilumab, whereas positive numbers indicate a net increase in per-patient costs with lenzilumab.
**Abbreviations:** CRP, C-reactive protein; mITT, modified intent-to-treat; SOC, standard of care.

The estimated clinical benefits and cost impact of adding lenzilumab to SOC at the cohort level are shown in **Table 3**. In the base case analysis, it was estimated that for a weekly cohort of 4,754 newly hospitalized patients, the addition of lenzilumab to SOC would result in 609 additional patients achieving SWOV, 599 IMV uses avoided, and 11,400 bed days, 12,978 ICU days, 15,831 IMV days, and 352 additional lives saved. Additionally, in this weekly cohort, lenzilumab plus SOC produced an estimated cost savings of £5,524,952 compared to SOC alone. At the cohort level the scenario analyses also resulted in estimated cost savings ranging from £606,442 (scenario #4) to £5,604,188 (scenario #1), with the exception of scenario #2 (full LIVE-AIR mITT population) which produced an estimated £27,304,581 in additional costs.

**Table 3.** Estimated clinical benefits and economic impact of lenzilumab plus SOC over SOC alone for a weekly cohort of newly hospitalized patients with COVID-19 in England for different population subgroups.

| Population<sup>a</sup> | Additional<br>Patients<br>Achieving<br>SWOV<sup>b</sup> | IMV Uses<br>Avoided<sup>b</sup> | Additional<br>Lives<br>Saved<sup>b</sup> | Bed Days<br>Saved<sup>b</sup> | ICU Days<br>Saved<sup>b</sup> | IMV Days<br>Saved<sup>b</sup> | Cost<br>Difference<br>Per Cohort<sup>c</sup> |
| --- | --- | --- | --- | --- | --- | --- | --- |
| **Base case: aged <85 years**<br>**with CRP <150 mg/L<sup>13,40,49,51</sup>**<br>**(n = 4,754)** | 609 | 599 | 352 | 11,400 | 12,978 | 15,831 | –£5,524,952 |
| **Scenario #1: aged <85 years**<br>**with CRP <150 mg/L,**<br>**receiving remdesivir<sup>13,40,49,51</sup>**<br>**(n = 1,792)** | 280 | 277 | 176 | 4,307 | 5,322 | 6,989 | –£5,604,188 |
| **Scenario #2: full LIVE-AIR**<br>**mITT population<sup>13,49</sup>**<br>**(n = 6,817)** | 443 | 443 | 293 | 6,726 | 8,249 | 12,611 | £27,304,581 |
| **Scenario #3: Black with CRP**<br>**<150 mg/L**<sup>13,40,49,51</sup><br>**(n = 200)** | 51 | 54 | 26 | 984 | 1,012 | 1,100 | −£1,995,391 |
| **Scenario #4: Black**<sup>13,49,51</sup><br>**(n = 256)** | 41 | 44 | 22 | 661 | 609 | 863 | −£606,442 |

**Notes:**
<sup>a</sup> Estimated cohort sizes were calculated using the total number of new COVID-19 hospital admissions in England over a one-week period (11,343; from January 17 to January 23, 2022),<sup>13</sup> adjusted to the population of interest based on SpO<sub>2</sub> <94% (60.1%),<sup>49</sup> CRP <150 mg/L (77.9%),<sup>40</sup> proportion <85 years (89.5%),<sup>51</sup> proportion receiving remdesivir for those with CRP <150 mg/L (37.7%),<sup>49</sup> and proportion Black (3.8%),<sup>51</sup> as appropriate.
<sup>b</sup> Values represent the difference of adding lenzilumab to SOC compared with SOC alone based on analyses of the LIVE-AIR trial.<sup>50</sup>
<sup>c</sup> Negative numbers indicate a net cost savings with lenzilumab, whereas positive numbers indicate a net budget impact with lenzilumab.
**Abbreviations:** COVID-19, Coronavirus Disease 19; CRP, C-reactive protein; ICU, intensive care unit; IMV, invasive mechanical ventilation; mITT, modified intent-to-treat; SOC, standard of care; SWOV, survival without ventilation.

### Sensitivity Analyses

Overall, the economic impact of the sensitivity analyses was variable relative to the base case results, with adjustments to the inputs for hospital resources costs having the greatest impact on the estimated results. The findings of the sensitivity analyses are summarized in **Table 4**.

**Table 4.**
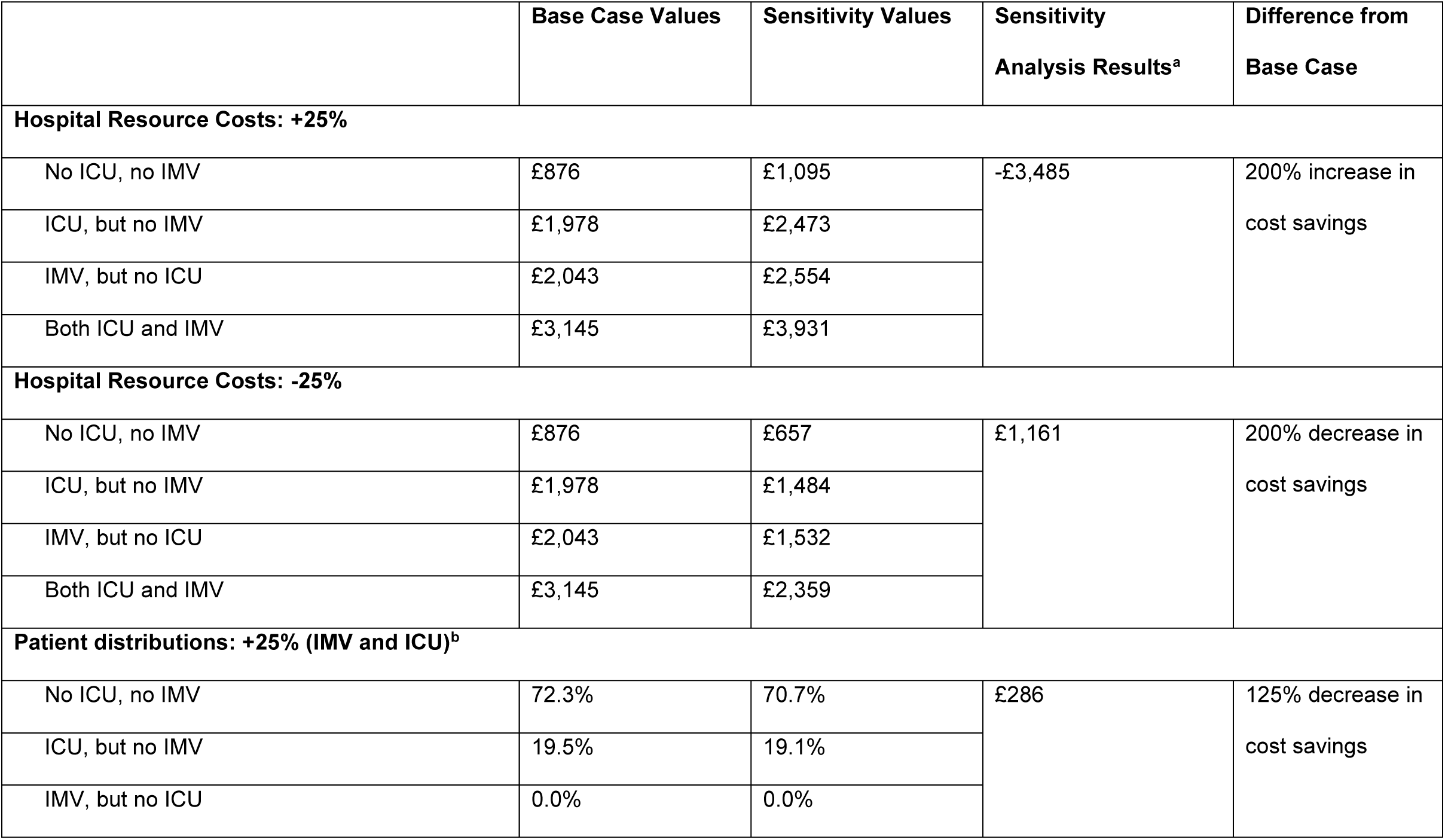
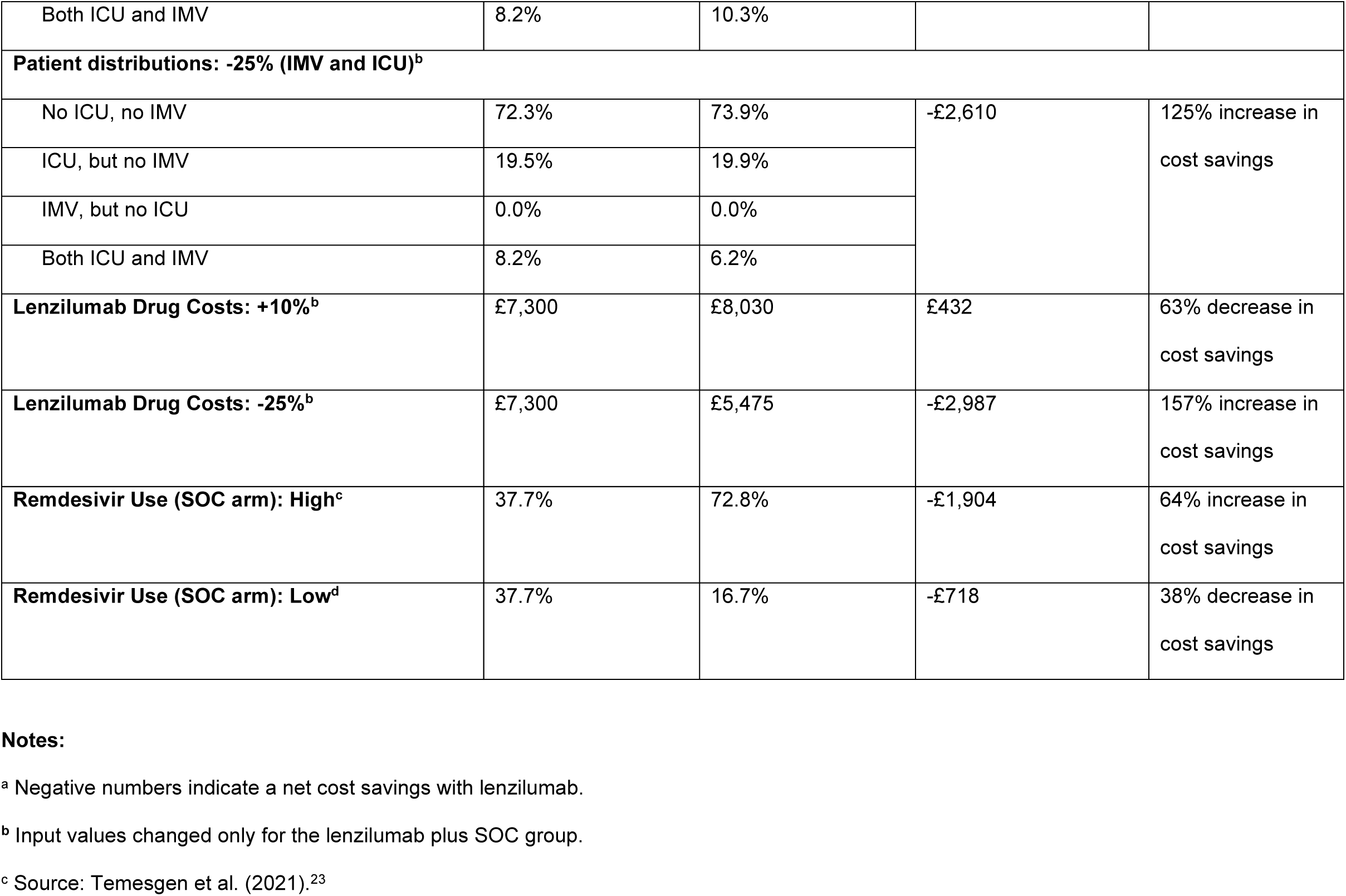
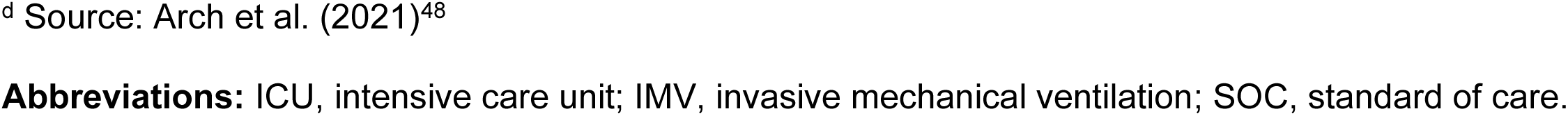
Sensitivity analyses.

Varying the hospital resource costs resulted in an additional cost of £1,161 per patient with lower (−25%) resource use costs and cost savings of £3,485 per patient with higher (+25%) resource use costs, a decrease and increase in cost savings of 200%, relative to the base case. Varying the patient distribution in the IMV and ICU level of care for the lenzilumab plus SOC arm resulted in a 125% change in cost savings relative to the base case; reducing the patients in the IMV and ICU level of care by 25% increased the cost savings to £2,610 per patient, while increasing the patients in the IMV and ICU level of care by 25% resulted in an additional cost of £286 per patient. Altering the lenzilumab drug costs produced cost savings of £2,987 per patient with lower drug costs (–25%) and additional costs of £432 per patient with higher drug costs (+10%), an 157% increase and 63% decrease in cost savings, respectively, relative to the base case, respectively. Finally, a higher use of remdesivir in the SOC arm resulted in cost savings of£1,904 per patient, an increase of 64% relative to the base. Conversely, a lower use of remdesivir in the SOC arm decreased the cost savings to £718 per patient, a reduction of 38%.

## Discussion

The present analysis evaluated the clinical and economic benefits of adding lenzilumab to SOC for the treatment of COVID-19 pneumonia from the NHS England perspective. The overall objective was to provide evidence to healthcare payers that may support the use of lenzilumab as a treatment option for this condition that urgently requires new efficacious therapies.

In the base case and all scenario analyses, treatment with lenzilumab plus SOC improved all specified clinical outcomes over SOC alone and resulted in a greater number of patients achieving SWOV and avoiding IMV use, as well as additional lives, bed days, ICU days, and IMV days saved. In the scenario analyses, the relative clinical benefits were smaller in the broader full mITT population scenario but greater in the narrower subgroup populations (ie, aged <85 years with CRP <150 mg/L receiving remdesivir and Black with CRP <150 mg/L, with or without remdesivir) compared to the base case.

Of the clinical outcomes assessed, SWOV is an important outcome for patients with COVID-19 from the NHS England perspective, as the average length of a patient’s hospital stay is greatly reduced by avoiding IMV.^59^ Notably, the average time to recovery for patients with IMV was more than double that of patients without IMV for all five analysis populations explored from the LIVE-AIR trial.^50^ In the base case analysis, addition of lenzilumab to SOC resulted in a 12.6% absolute reduction in the probability of requiring IMV compared with SOC alone among patients aged <85 years with CRP <150 mg/L. As observed in the LIVE-AIR trial, the estimates for failure to achieve SWOV were similar between patients with CRP <150 mg/L regardless of age and patients aged <85 years with CRP <150 mg/L (hazard ratio [95% confidence interval] of 2.54 [1.46-4.41] and 3.04 [1.68-5.51], respectively).^39,40^ These results suggest that the clinical benefits observed in the present base case analysis would also likely be generalizable to a broader population of patients with CRP <150 mg/L, irrespective of age.

In the base case analysis, lenzilumab was associated with per-patient cost savings from the NHS England perspective. Although there were increases in drug acquisition and administration costs associated with the addition of lenzilumab to SOC compared with SOC alone, these additional costs were offset by the reduced costs for hospital resource use among patients treated with lenzilumab. This resulted in a net per-patient cost savings of £1,162, and an estimated total savings of £5,524,952 for a weekly cohort of 4,754 newly hospitalized patients with CRP <150 mg/L and aged <85 years. Results from the sensitivity analyses suggested that the variable with the greatest impact on the estimated cost savings of adding lenzilumab to SOC versus SOC alone was hospital resource costs (±25%), resulting in a change of 200% from the base case analysis. By comparison, varying patient distributions to different levels of care (±25%), lenzilumab drug costs (+10% and –25%) and using different assumptions for remdesivir use in the SOC alone arm (high estimate of 72.8%, low estimate of 16.7%) had less of an impact on the cost savings associated with adding lenzilumab to SOC in the base case analysis.

In addition to the clinical benefits observed in the various populations examined, the results show that adding lenzilumab to SOC for a narrower group of patients with baseline CRP <150 mg/L, aged <85 years, and receiving remdesivir may also provide economic benefits for UK hospitals, with a net per-patient cost savings of £3,127. However, in a broader population of patients with COVID-19 pneumonia without consideration of CRP criteria or age, adding lenzilumab to SOC had additional per-patient costs of £4,005, suggesting that there is greater economic value in providing lenzilumab to a more targeted population.

Evidence suggests that CRP levels at the time of admission have a positive correlation with disease severity and are associated with adverse clinical outcomes in patients hospitalized with COVID-19.^23,34-37,40^ Furthermore, one of the strongest independent predictors of critical illness is CRP levels >100 mg/L at the time of admission^34,35^, with patients having CRP levels >150 mg/L considered to be high risk for escalation of respiratory support (ie, need for non-invasive ventilation or intubation) or death.^60^ Given that in patients aged <85 years with CRP levels <150 mg/L, irrespective of remdesivir use, lenzilumab produced favorable clinical outcomes and cost savings, it appears that using lenzilumab as an early intervention would be most effective and provide the best economic value. As testing for CRP levels is accessible and inexpensive in the UK hospital setting and has previously been recommended by the National Institute for Health and Care Excellence (NICE) as an option to inform the diagnosis and treatment of pneumonia in adults,^61^ it may be a valuable and feasible approach to identify hospitalized COVID-19 patients who may benefit most from lenzilumab treatment.

Clinical trials frequently underrepresent the Black population, despite this population being disproportionately affected by COVID-19.^62^ Consequently, there is limited evidence across racial populations regarding differences in disease severity, outcomes, and treatments.^62^ An exploratory analysis of the LIVE-AIR trial revealed that Black and African American patients, particularly those with a CRP level <150 mg/L, demonstrated the greatest response to treatment with lenzilumab.^41^ In the current analysis, the use of lenzilumab in Black patients with CRP <150 mg/L resulted in cost savings of £9,977 per patient, and a cost savings of £2,369 per patient when all Black patients were assessed. Assuming the Black and African American subgroup of LIVE-AIR are representative of potential outcomes in the Black African and Black Caribbean persons in the UK, these findings may be significant since this demographic is disproportionally affected by COVID-19, with an increased rate of death.^42^

As with extrapolation of any clinical trial results, this study had several limitations. The analysis used data from the LIVE-AIR Phase 3 clinical trial that may have been subject to selection bias and that was predominantly conducted in US hospitals. Consequently, the findings from the trial may not be fully generalizable to a UK population and/or UK hospital setting based on potential differences in treatment patterns (eg, length of hospital stay) for hospitalized patients with COVID-19 between the US and UK. It should also be noted that the LIVE-AIR trial was conducted earlier in the pandemic and thus, prior to vaccinations and the emergence of new COVID-19 variants, both of which have been reported to affect hospital length of stay.^11,63,64^ As a result, time to recovery inputs derived from the clinical trial may differ from current real-world values. However, it is anticipated that the ongoing ACTIV-5/BET-B trial, which has completed the enrollment of over 400 patients in the primary analysis population, will provide data on the use of lenzilumab for hospitalized patients infected with different COVID-19 variants and with differing vaccination statuses.^46,47^ Additionally, the Black subgroups from the LIVE-AIR trial that were assessed in the per-patient calculator were limited by small sample sizes and will require further validation using the results from the upcoming ACTIV-5/BET-B trial. Consideration should also be given to the fact that the current analyses were conducted *ex ante* and therefore, were conducted prior to regulatory approval of lenzilumab. Notably, a Conditional Marketing Authorisation (CMA) request for lenzilumab was submitted to the Medicines and Healthcare products Regulatory Agency (MHRA) on June 11, 2021 and is currently under review. Although the US Food and Drug Administration (FDA) (September 9, 2021) initially declined the request for emergency use authorization (EUA) of lenzilumab for the treatment of newly hospitalized COVID-19 patients based on the LIVE-AIR trial data, concluding further clinical data are needed, the FDA has invited Humanigen to submit any supplemental data as they become available.^65^. It is anticipated that the ongoing ACTIV-5/BET-B trial may provide additional efficacy and safety data sufficient to further support the use of lenzilumab for the treatment of hospitalized COVID-19 patients with a CRP <150 mg/L.^46,47^

Although approximately 83.9% and 64.3% of individuals ≥12 years of age in the UK are now double and triple vaccinated, respectively, as of January 25, 2022,^9^ there remains an ongoing need for effective treatments for patients hospitalized with COVID-19 with the emergence of new variants.^66^ As per the NICE COVID-19 Treatment Guidelines, the current mainstays of care for hospitalized patients include remdesivir, corticosteroids, or both, casirivimab with imdevimab, and tocilizumab.^67^ However, these therapies may have a greater impact early in the disease course (remdesivir)^68^ or in patients with advanced disease (corticosteroids and tocilizumab).^69,70^ Therefore, effective treatment options are limited for hospitalized and hypoxic patients with COVID-19 pneumonia, particularly early in the disease process to prevent ICU admission, IMV, and/or death. Lenzilumab is a particularly promising therapeutic as it addresses an unmet need for treatment options that prevent the progression to IMV and/or death in hospitalized patients. Additionally, the mechanism of action of lenzilumab functions by binding and neutralizing GM-CSF, which is produced in response to COVID-19 infection. Therefore, as was reported in the COVID-19 Therapeutics Strategy published by the European Commission, its efficacy is not anticipated to be affected by the new COVID-19 variants, including Omicron.^38^

## Conclusions

Overall, the findings from this analysis support the clinical benefits for SWOV, ventilator use, time to recovery, mortality, time in ICU, and time on IMV, as well as an economic benefit from the NHS England perspective associated with adding lenzilumab to SOC for patients with COVID-19 pneumonia. Lenzilumab provides clinical benefits to a broad population of patients with characteristics similar to the full mITT study cohort from the LIVE-AIR trial. Importantly, when used in patients aged <85 years with CRP <150 mg/L, lenzilumab appears to be particularly effective and is estimated to result in cost savings to the UK hospitals. Lenzilumab treatment in the Black population also resulted in clinical benefits and cost savings. As Black persons are hyper-vulnerable to COVID-19,^42^ this is a critical finding. Further clinical benefits and cost savings were observed with concomitant use of remdesivir with lenzilumab. These results support the use of lenzilumab as a standard treatment option for COVID-19 pneumonia and may help with its consideration for adoption in the hospital setting by hospital formulary decisions makers should an EUA or full approval through a Biologics License Application be granted.

## Data Availability

All data produced in the present work are contained in the manuscript.

## Abbreviations

AE: adverse event
CMA: Conditional Marketing Authorisation
COVID-19: Coronavirus Disease 2019
CPI: consumer price inflation
CRP: C-reactive protein
EUA: emergency use authorization
EUR: euros
FDA: Food and Drug Administration
GBP: pound sterling
GM-CSF: granulocyte-macrophage colony-stimulating factor
ICU: intensive care unit
IL-6: interleukin-6
IMV: invasive mechanical ventilation
MHRA: Medicines and Healthcare products Regulatory Agency
mITT: modified intent-to-treat
NHS: National Health Service
NICE: National Institute for Health and Care Excellence
NNT: number needed to treat
ONS: Office of National Statistics
SOC: standard of care
SpO_2_: oxygen saturation
SWOV: survival without ventilation
UK: United Kingdom
US: United States.

## Ethics Approval and Informed Consent

The current budget impact model did not require IRB oversight or approval. The LIVE-AIR trial on which the clinical efficacy was based was conducted in accordance with the Good Clinical Practice guidelines of the International Council for Harmonization E6 and the principles of the Declaration of Helsinki. The protocol was approved by Advarra, the central institutional review board. The ID number was Pro00043147. Additionally, local ethics committees at Mayo Clinic and the University of Southern California approved the protocol. Participants provided written informed consent.

## Acknowledgments

The authors acknowledge Cameron Durant (Humanigen Inc.) and Avery Hughes (EVERSANA) for their specific contributions to this project.

## Author Contributions

All authors made substantial contributions to conception and design, acquisition of data, or analysis and interpretation of data; took part in drafting the article or revising it critically for important intellectual content; agreed to submit to the current journal; gave final approval of the version to be published; and agree to be accountable for all aspects of the work

## Footnotes

a SWOV is a robust composite endpoint used in many of the recent COVID-19 studies that is less prone to favor treatments with discordant effects on survival and days free of ventilation while avoiding the need for sample sizes approaching those of mortality trials to enable timely availability of study results.

b The mITT population was the analysis set used for the primary analysis of efficacy, defined as all randomized subjects who received at least one dose of study drug under the documented supervision of the principal investigator or sub-investigator and excluding sites that experienced documented limitations to access of basic supportive care for COVID-19.

c Time to recovery was a pre-specified secondary outcome of the LIVE-AIR trial and was defined as the first day on which a patient was discharged or ready for discharge by satisfying one of the following 3 categories from the 8-point ordinal scale: hospitalized, not requiring supplemental oxygen, no longer requiring ongoing medical care; not hospitalized, limitation on activities and/or requiring home oxygen; not hospitalized, no limitations on activities.

