## Supplemental Appendix for "Clinical and economic benefits of lenzilumab plus standard of care compared with standard of care alone for the treatment of hospitalized patients with Coronavirus Disease 19 (COVID-19) from the perspective of National Health Service England"

### Supplementary Calculations

#### *Treatment Efficacy*

The LIVE-AIR trial was used to inform the calculator inputs for treatment efficacy of lenzilumab plus SOC compared to SOC alone.[1] As previously reported in the US hospital perspective analysis by Kilcoyne and colleagues (2022),[2] treatment efficacy inputs for the base case and scenario analyses included failure to achieve SWOV, mortality, time to recovery, time in ICU, and time on IMV. All treatment efficacy input values used in the model are provided in **Supplementary Table 1**.

#### *Drug Acquisition and Administration Costs*

Lenzilumab is administered as three 1-hour intravenous infusions, each administered 8 hours apart.[3] Administration costs for lenzilumab were estimated using a time and motion study by Burcombe and colleagues,[4] which reported that the aseptic preparation time of the monoclonal antibody trastuzumab was 34.5 minutes. Preparation time using aseptic technique was used for lenzilumab administration as its preparation requires the compounding of six 100 mg vials and therefore is considered a medium-risk for contamination.[5] A pharmacist specialist hourly wage of £57.00 was reported by the National Institute for Health and Care Excellence (NICE) review for ravulizumab.[6] This cost was then inflated from 2019 GBP to 2021 GBP to provide the estimated hourly wage of £58.99 per hour.[7] The product of the inflated wage and the preparation time resulted in an estimated administration cost of £33.92 for each infusion of lenzilumab, and a total administration cost of £101.76 for the full course of lenzilumab treatment.

Remdesivir is administered as five intravenous infusions, each administered one day apart.[8] The first infusion is a loading dose of 200 mg and all subsequent infusions are maintenance doses of 100 mg.[8] Administration costs for remdesivir were estimated based on the NICE review for ravulizumab, which reported a preparation time of 15 minutes without aseptic technique.[6] It was assumed that additional time for aseptic compounding was not needed for remdesivir as its preparation includes no more than two drug vials and thus is considered a low-risk for contamination.[5] The same inflated pharmacist specialist hourly wage of £58.99 was assumed,[7] and the product of the inflated wage and the preparation time resulted in an estimated administration cost of £14.75 per infusion. The total administration cost for the full course of remdesivir treatment was estimated to be £73.75.

Subsequently, the total administration cost for the full course of remdesivir treatment was then multiplied by the proportion of patients receiving remdesivir in each treatment arm. With the exception of scenario #1 (aged <85 years with CRP <150 mg/L and receiving remdesivir), it was assumed that 72.1% of lenzilumab plus SOC patients would also receive remdesivir, in alignment with the actual use in the LIVE-AIR trial.[3] However, as scenario #1 includes only patients aged <85 years with CRP <150 mg/L and receiving remdesivir, remdesivir use was set at 100% for both the lenzilumab plus SOC and SOC alone arms. Based on the physician survey for the treatment of patients with CRP <150 mg/L, 37.7% of patients in the SOC alone arm were assumed to receive remdesivir in the base case analysis and scenario #3 (Black with CRP <150 mg/L).[9] Finally, 48.4% of patients in the SOC alone arm were assumed to receive remdesivir in scenarios #2 (full mITT population) and #4 (Black patients from the full mITT population), as per the physician survey for the treatment of patients with $SpO_2$ ≤94%, regardless of CRP level.[9] The calculations for remdesivir acquisition and administration costs are presented in **Supplementary Table 2**.

The total drug administration costs within each treatment arm were then calculated as the sum of the lenzilumab administration costs and the remdesivir administration costs. As there were no lenzilumab administration costs in the SOC alone arm, the total drug administration costs were equivalent to the remdesivir administration costs. Based on the balanced use of corticosteroids in both arms of the LIVE-AIR trial,[3] it was assumed that lenzilumab would not impact the utilization or cost of background therapies. For this reason, costs associated with other SOC drugs were not included as separate inputs in the calculator. The calculations for the total drug administration costs are presented in **Supplementary Table 3.**

#### *Hospital Resource Costs*

Hospital resource costs for each level of care (no ICU, no IMV; ICU, but no IMV; IMV, but no ICU; both ICU and IMV) were determined in three steps. First, the daily hospital resource cost inputs were estimated based on a previous healthcare cost model for COVID-19.[10] Second, time to recovery results from the LIVE-AIR trial were applied to estimate the average total hospital resource cost for each level of care.[1] Finally, the weighted average hospital resource cost per patient was calculated by applying the patient distributions within the four levels of care observed in the LIVE-AIR trial.[1] With the exception of the lenzilumab and remdesivir acquisition and administration (ie, pharmacy labor) costs, it was assumed that these hospital resource costs would capture all costs incurred by the NHS during the index hospitalization. The three steps used to calculate the hospital resource costs are described in additional detail in the subsequent sections. All hospital resource costs and use inputs are presented in **Supplementary Table 4**.

##### Step 1: Daily Hospital Resource Costs by Level of Care

Daily hospital resource costs by level of care were estimated using a previously published healthcare cost model that evaluated the costs of the COVID-19 pandemic associated with obesity in Europe by Czernichow and colleagues.[10] The study used country-specific data on the number of hospitalizations from January 1, 2020 to June 30, 2020 within the model and was selected as it provides data on the actual cost per bed type per day incurred, which was deemed more relevant for the NHS perspective than hospital charges, which are more commonly reported.

Supplementary Table 1 from Czernichow and colleagues[10] reports the mean cost per bed type per day in the UK; it was assumed that these costs incorporated all costs associated with SOC in the UK, including corticosteroid use. As a cost corresponding to the "IMV, but no ICU" level of care was not provided in the study, it was calculated as the difference between the "both ICU and IMV" and "ICU, but not IMV" groups, added to the "no ICU, no IMV" cost. Czernichow and colleagues reported all costs in their study in EUR, using an exchange rate of 1.11 EUR to 1 GBP.[10] Using this exchange rate, costs were reconverted to GBP and then inflated from 2020 to 2021 values.[7] The converted and inflated daily hospital resource costs by level of care are presented in **Supplementary Table 5**.

#### Step 2: Total Hospital Resource Cost for Lenzilumab Plus SOC and SOC Alone by Level of Care

To obtain the average total hospital resource costs for a patient receiving each level of care within each treatment arm, the mean daily hospital resource costs calculated in Step 1 were multiplied by the average time to recovery for the corresponding level of care (**Supplementary Table 4**) derived from the LIVE-AIR trial.[1] Due to the differences in time to recovery between the treatment arms,[1] total hospital resource costs were calculated separately for lenzilumab plus SOC and SOC alone arms. Mean daily hospital resource costs, time to recovery data, and calculated average total hospital resource costs by level of care for the case analysis are presented in **Supplementary Table 6**.

#### Step 3: Weighted Average Hospital Resource Costs per Patient for Lenzilumab Plus SOC and SOC Alone

The weighted average hospital resource cost per hospitalized patient was calculated as the sum product of total hospital resource costs by level of care calculated in Step 2 and the corresponding percentage of patients within each level of care (**Supplementary Table 4**) from the LIVE-AIR trial.[1] As a result of the differences in total hospital resource costs and patient distribution for each treatment arm in the LIVE-AIR trial,[1] separate calculations were performed for lenzilumab plus SOC and SOC alone. The corresponding values for the base case analysis are presented in **Supplementary Table 7**.

**Supplementary Table 1** Treatment efficacy model inputs from LIVE-AIR trial data.[1,a]

| | **Failure to achieve SWOV[b]** | **Mortality[b]** | **Time in ICU (days)[c]** | **Time on IMV (days)[c]** |
|---|---|---|---|---|
| **Base case: aged <85 years with CRP <150 mg/L** | | | | |
| Lenzilumab plus SOC (*n* = 159) | 8.4% (5.0-14.0) | 6.5% (3.5-11.7) | 3.48 (7.8) | 1.92 (21.2) |
| SOC alone (*n* = 178) | 21.2% (15.9-28.1) | 13.9% (9.6-20.9) | 6.21 (10.6) | 5.25 (17.5) |
| **Scenario #1: aged <85 years with CRP <150 mg/L, receiving remdesivir** | | | | |
| Lenzilumab plus SOC (*n* = 123) | 9.2% (5.2-15.9) | 7.5% (4.0-13.9) | 3.79 (8.2) | 2.12 (20.7) |
| SOC alone (*n* = 131) | 24.8% (18.2-33.2) | 17.3% (11.7-25.1) | 6.76 (11.2) | 6.02 (16.9) |
| **Scenario #2: full LIVE-AIR mITT population** | | | | |
| Lenzilumab plus SOC (*n* = 236) | 15.6% (11.5-20.9) | 9.5% (6.4-14.1) | 5.40 (9.6) | 3.53 (19.2) |

| | | | | |
|---|---|---|---|---|
| SOC alone (*n* = 243) | 22.1% (17.4-27.9) | 13.9% (10.1-19.0) | 6.61 (10.7) | 5.38 (17.5) |
| **Scenario #3: Black with CRP <150 mg/L** | | | | |
| Lenzilumab plus SOC (*n* = 25) | 4.0% (0.6-25.2) | 4.0% (0.6-25.6) | 1.48 (5.7) | 1.12 (22.4) |
| SOC alone (*n* = 26) | 29.3% (15.1-51.9) | 17.1% (6.8-39.4) | 6.54 (11.1) | 6.62 (16.6) |
| **Scenario #4: Black** | | | | |
| Lenzilumab plus SOC (*n* = 38) | 13.2% (5.7-28.8) | 7.9% (2.6-22.5) | 4.68 (9.8) | 3.11 (19.7) |
| SOC alone (*n* = 33) | 29.1% (16.3-48.5) | 16.5% (7.2-35.2) | 7.06 (10.9) | 6.48 (16.8) |

**Notes:**

[a] All data were censored at 28 days following trial enrollment. Data are presented for the mITT population.

[b] These values were derived from Kaplan-Meier analysis. 95% confidence intervals are reported in brackets.

[c] Values reported are means with standard deviations.

**Abbreviations:** CRP, C-reactive protein; ICU, intensive care unit; IMV, invasive mechanical ventilation; mITT, modified intent-to-treat; SOC, standard of care; SWOV, survival without ventilation.

**Supplementary Table 2** Calculations for remdesivir acquisition and administration costs.

| Scenario | Remdesivir Use | | Remdesivir Acquisition Costs | | Remdesivir Administration Costs | |
|---|---|---|---|---|---|---|
| | **Lenzilumab + SOC[a]** | **SOC Alone[b]** | **Lenzilumab + SOC** | **SOC Alone** | **Lenzilumab + SOC** | **SOC Alone** |
| | **A** | **B** | **C = A × £2,040[c]** | **D = B × £2,040[c]** | **E = A × £73.75[d]** | **F = B × £73.75[d]** |
| **Base case: aged <85 years with CRP < 150 mg/L** | 72.1% | 37.7% | £1,470.84 | £769.08 | £53.17 | £27.80 |
| **Scenario #1: aged <85 years with CRP <150 mg/L, receiving remdesivir** | 100.0% | 100.0% | £2,040.00 | £2,040.00 | £73.75 | £73.75 |
| **Scenario #2: full LIVE-AIR mITT population** | 72.1% | 48.4% | £1,470.84 | £987.36 | £53.17 | £35.70 |
| **Scenario #3: Black with CRP <150 mg/L** | 72.1% | 37.7% | £1,470.84 | £769.08 | £53.17 | £27.80 |
| **Scenario #4: Black** | 72.1% | 48.4% | £1,470.84 | £987.36 | £53.17 | £35.70 |

**Notes:**

[a] With the exception of scenario #1, it was assumed that 72.1% of lenzilumab plus SOC patients received concomitant remdesivir, in alignment with the actual use in the LIVE-AIR trial.[3] As scenario #1 considered patients aged <85 years with CRP <150 mg/L and receiving remdesivir, remdesivir use was set at 100.0%.

[b] Based on the physician survey for the treatment of patients with CRP <150 mg/L, remdesivir use was estimated at 37.7% for patients who received SOC alone in the base case and scenario #3 (Black with CRP <150 mg/L).[9] Remdesivir use was set at 100.0% for scenario #1 as it only included patients aged <85 years with CRP <150 mg/L and receiving remdesivir. For scenarios #2 (full mITT population) and #4 (Black patients from the full mITT population), 48.4% of patients in the SOC alone arm were assumed to receive remdesivir, based on the physician survey for the treatment of patients with $SpO_2$ ≤94%, regardless of CRP level.[9]

[c] This cost was calculated using a cost of £340 per 100 mg vial[11] and assuming a course of 6 vials based on the recommended 200 mg loading dose on day 1 and 100 mg maintenance doses on days 2-5.[8]

[d] Value calculated based on an assumed administration cost of £14.75 per infusion of remdesivir, as per the pharmacy labor cost associated with the administration of a monoclonal antibody, with no aseptic preparation.[6] With five infusions per treatment course, this resulted in a total administration cost of £73.75 for a full course of remdesivir treatment. For additional details, see the **Drug Acquisition and Administration Costs** section.

**Abbreviations:** CRP, C-reactive protein; mITT, modified intent-to-treat; SOC, standard of care; $SpO_2$, oxygen saturation.

**Supplementary Table 3** Calculations for total drug administration costs.

| **Scenario** | **Lenzilumab Administration Costs** | | **Remdesivir Administration Costs** | | **Total Drug Administration Costs** | |
|---|---|---|---|---|---|---|
| | **Lenzilumab + SOC[a]** | **SOC Alone** | **Lenzilumab + SOC** | **SOC Alone** | **Lenzilumab + SOC** | **SOC Alone** |
| | A | B | C | D | **E = A + C** | **F = B + D** |
| **Base case: aged <85 years with CRP < 150 mg/L** | £101.76 | £0.00 | £53.17 | £27.80 | **£154.93** | **£27.80** |
| **Scenario #1: aged <85 years with CRP <150 mg/L, receiving remdesivir** | £101.76 | £0.00 | £73.75 | £73.75 | **£175.51** | **£73.75** |
| **Scenario #2: full LIVE-AIR mITT population** | £101.76 | £0.00 | £53.17 | £35.70 | **£154.93** | **£35.70** |
| **Scenario #3: Black with CRP <150 mg/L** | £101.76 | £0.00 | £53.17 | £27.80 | **£154.93** | **£27.80** |
| **Scenario #4: Black** | £101.76 | £0.00 | £53.17 | £35.70 | **£154.93** | **£35.70** |

**Note:**

[a] Value was calculated assuming a cost of £33.92 per intravenous infusion of lenzilumab, based on the pharmacy labor cost associated with the aseptic preparation and administration of a monoclonal antibody.[4,6] With three infusions per treatment course, this resulted in a total administration cost of £101.76 for a full course of lenzilumab treatment. For additional details, see the **Drug Acquisition and Administration Costs** section.

**Abbreviations:** CRP, C-reactive protein; mITT, modified intent-to-treat; SOC, standard of care.

**Supplementary Table 4** Hospital resource costs and use inputs from LIVE-AIR trial data.[a]

| | **No ICU, no IMV** | **ICU, but no IMV** | **IMV, but no ICU[b]** | **Both ICU and IMV** |
|---|---|---|---|---|
| **Hospital Resource Costs[10]** | | | | |
| Cost per day | £876 | £1,978 | £2,043[c] | £3,145 |
| **Patient distributions[1]** | | | | |
| **Base case: aged <85 years with CRP <150 mg/L** | | | | |
| Lenzilumab plus SOC (*n* = 159) | 72.3% | 19.5% | 0.0% | 8.2% |
| SOC alone (*n* = 178) | 60.1% | 19.1% | 0.6% | 20.2% |
| **Scenario #1: aged <85 years with CRP <150 mg/L, receiving remdesivir** | | | | |
| Lenzilumab plus SOC (*n* = 123) | 72.4% | 18.7% | 0.0% | 8.9% |
| SOC alone (*n* = 131) | 62.6% | 13.0% | 0.8% | 23.7% |
| **Scenario #2: full LIVE-AIR mITT population** | | | | |
| Lenzilumab plus SOC (*n* = 236) | 64.4% | 20.3% | 0.0% | 15.3% |
| SOC alone (*n* = 243) | 58.0% | 20.2% | 0.4% | 21.4% |
| **Scenario #3: Black with CRP <150 mg/L** | | | | |
| Lenzilumab plus SOC (*n* = 25) | 84.0% | 12.0% | 0.0% | 4.0% |
| SOC alone (*n* = 26) | 57.7% | 11.5% | 3.9% | 26.9% |

| | | | | |
|---|---|---|---|---|
| **Scenario #4: Black** | | | | |
| Lenzilumab plus SOC (*n* = 38) | 71.1% | 15.8% | 0.0% | 13.2% |
| SOC alone (*n* = 33) | 54.6% | 15.2% | 3.0% | 27.3% |
| **Time to recovery (days)[1]** | | | | |
| **Base case: aged <85 years with CRP <150 mg/L** | | | | |
| Lenzilumab plus SOC (*n* = 159) | 6.49 | 12.43 | 16.61 | 25.54 |
| SOC alone (*n* = 178) | 7.07 | 8.82 | 17.93 | 27.56 |
| **Scenario #1: aged <85 years with CRP <150 mg/L, receiving remdesivir** | | | | |
| Lenzilumab plus SOC (*n* = 123) | 7.09 | 13.68 | 16.32 | 25.09 |
| SOC alone (*n* = 131) | 7.04 | 9.94 | 17.88 | 27.48 |
| **Scenario #2: full LIVE-AIR mITT population** | | | | |
| Lenzilumab plus SOC (*n* = 236) | 6.85 | 12.87 | 17.29 | 26.58 |
| SOC alone (*n* = 243) | 7.47 | 10.00 | 17.19 | 26.43 |
| **Scenario #3: Black with CRP <150 mg/L** | | | | |
| Lenzilumab plus SOC (*n* = 25) | 6.57 | 14.00 | 18.22 | 28.00 |
| SOC alone (*n* = 26) | 7.47 | 10.00 | 17.19 | 26.43 |
| **Scenario #4: Black** | | | | |
| Lenzilumab plus SOC (*n* = 38) | 6.85 | 14.67 | 18.22 | 28.00 |

| | | | | |
|---|---|---|---|---|
| SOC alone (*n* = 33) | 7.00 | 13.40 | 16.91 | 26.00 |

**Notes:**

[a] All data were censored at 28 days following trial enrollment. Data are presented for mITT population.

[b] In the absence of time to recovery data for the “IMV, but no ICU” group from the LIVE-AIR trial, this was calculated as the time to recovery for the “both ICU and IMV” group multiplied by (12.1/18.6), the ratio for mean length of stay between “IMV, but no ICU” and “both ICU and IMV”, respectively from Di Fusco et al.[12]

[c] This cost was calculated as the difference between the “Both ICU and IMV” and “ICU, but not IMV” groups, added to the “no ICU, no IMV” cost.

**Abbreviations:** CRP, C-reactive protein; ICU, intensive care unit; IMV, invasive mechanical ventilation; mITT, modified intent-to-treat; SOC, standard of care.

**Supplementary Table 5** Conversion and inflation of daily hospital resource costs.

| **Level of Care** | **Mean Cost Per Bed Type (EUR 2020)[10]** | **Mean Cost Per Bed Type (GBP 2020)[10]** | **Mean Cost Per Bed Type (GBP 2021)[7]** |
|---|---|---|---|
| | **A** | **B = A/1.11** | |
| **No ICU, no IMV** | €961 | £866 | **£876** |
| **ICU, but no IMV** | €2,170 | £1,955 | **£1,978** |
| **IMV, but no ICU** | €2,241[a] | £2,019 | **£2,043** |
| **Both ICU and IMV** | €3,450 | £3,108 | **£3,145** |

**Note:**

[a] This cost was calculated as the difference between the "Both ICU and IMV" and "ICU, but not IMV" groups, added to the "no ICU, no IMV" cost.

**Abbreviations:** EUR, euros; GBP, pound sterling; ICU, intensive care unit; IMV, invasive mechanical ventilation.

**Supplementary Table 6** Base case calculations for total hospital resource cost by treatment arm.

| **Level of Care** | **Mean Daily Hospital Resource Cost** | **Time to Recovery (days)[1]** | | **Total Hospital Resource Costs** | |
|---|---|---|---|---|---|
| | | **Lenzilumab + SOC** | **SOC Alone** | **Lenzilumab + SOC** | **SOC Alone** |
| | A | B | C | **D = A × B** | **E = A × C** |
| **No ICU, no IMV** | £876 | 6.49 | 7.07 | **£5,687** | **£6,196** |
| **ICU, but no IMV** | £1,978 | 12.43 | 8.82 | **£24,590** | **£17,448** |
| **IMV, but no ICU** | £2,043 | 16.61 | 17.93 | **£33,935** | **£36,632** |
| **Both ICU and IMV** | £3,145 | 25.54 | 27.56 | **£80,323** | **£86,676** |

**Abbreviations:** ICU, intensive care unit; IMV, invasive mechanical ventilation; SOC, standard of care.

**Supplementary Table 7** Base case calculations of the weighted average total hospital resource cost by treatment arm.

| Level of Care | Total Hospital Resource Costs | | Patient Distribution[1] | | Weighted Total Hospital Resource Costs[a] | |
|---|---|---|---|---|---|---|
| | Lenzilumab + SOC | SOC Alone | Lenzilumab + SOC | SOC Alone | Lenzilumab + SOC | SOC Alone |
| | A | B | C | D | **E = A × C** | **F = B × D** |
| **No ICU, no IMV** | £5,687 | £6,196 | 72.3% | 60.1% | **£4,112** | **£3,724** |
| **ICU, but no IMV** | £24,590 | £17,448 | 19.5% | 19.1% | **£4,795** | **£3,335** |
| **IMV, but no ICU** | £33,935 | £36,632 | 0.0% | 0.6% | **£0** | **£220** |
| **Both ICU and IMV** | £80,323 | £86,676 | 8.2% | 20.2% | **£6,586** | **£17,509** |
| **Weighted Average Total Hospital Resource Costs per Hospitalized Patient** | | | | | **£15,493** | **£24,784** |

**Note:**

[a] Rounded values are presented throughout the table; however, unrounded values were used to calculate the weighted average total hospital resource costs per hospitalized patient in the cost calculator.

**Abbreviations:** ICU, intensive care unit; IMV, invasive mechanical ventilation; SOC, standard of care.
